# Psychosocial factors at work and fertility and menstrual disorders: a systematic review

**DOI:** 10.1101/2022.06.17.22276473

**Authors:** Natsu Sasaki, Kotaro Imamura, Kazuhiro Watanabe, Yui Hidaka, Asuka Sakuraya, Emiko Ando, Hisashi Eguchi, Akiomi Inoue, Kanami Tsuno, Yu Komase, Mako Iida, Yasumasa Otsuka, Mai Iwanaga, Yuka Kobayashi, Reiko Inoue, Akihito Shimazu, Akizumi Tsutsumi, Norito Kawakami

## Abstract

**Objective:** This systematic review aimed to evaluate the association between psychosocial factors at work and menstrual abnormalities or fertility by collecting the literature that had utilized a longitudinal or prospective cohort design.

**Data sources:** MEDLINE, EMBASE, PsycINFO, PsycARTICLES, and Japan Medical Abstracts Society electronic databases were searched for published studies from inception to 26th February 2020.

**Study eligibility criteria:** The inclusion criteria for this systematic review were defined as follows: (P) Adult female workers (over 18 years old), (E) Presence of adverse psychosocial factors at work, (C) Absence of adverse psychosocial factors at work, and (O) Any menstrual cycle disorders, menstrual-related symptoms, or fertility. The study design was limited to prospective/longitudinal studies.

**Study appraisal and synthesis methods:** The included studies were descriptively summarized in a narrative format.

**Results:** Database searching yielded 12,868 abstracts. After the screening process, six studies were included. The outcomes were fertility (n=3), early menopause (n=1), endometriosis (n=1), and serum hormones (n=1). Three included studies presented significant findings: women with high job demands were significantly less likely to conceive; working over 40 hours per week and lifting or moving a heavy load >15 times per day significantly increased the duration of pregnancy attempts; women with rotating night shift work had increased risk of earlier menopause. All the study (n=3) that examined the association of night shift/rotating work with fertility outcomes showed no significant difference. No study investigated the association of psychosocial factors at work with menstrual abnormality.

**Conclusion:** This review revealed insufficient high-level evidence on the association of psychosocial factors at work with fertility and menstrual disorders. Future well-designed studies are needed.

**Trial registration:** The study protocol was registered at the UMIN registry (registration number: UMIN000039488). The registration date is 14th Feb 2020.

URL: https://upload.umin.ac.jp/cgi-bin/ctr/ctr_view_reg.cgi?recptno=R000044704

**Review protocol:** Protocol paper is available in preprint format.

URL:https://assets.researchsquare.com/files/rs-179301/v1/48845b10-5ec4-4d48-8918-3dcf0e0edded.pdf?c=1631873381

**Condensation:** This systematic review evaluated the association between psychosocial factors at work and fertility and menstrual disorders.

**A. Why was this study conducted?**

- The longitudinal studies which examined the associations between psychosocial factors at work and menstrual abnormalities or fertility have not been systematically presented yet.

**B. What are the key findings?**

- The outcomes were fertility (n=3), early menopause (n=1), endometriosis (n=1), and serum hormones (n=1), in the included six studies.
- All the study (n=3) that examined the association of night shift/rotating work with fertility outcomes showed no significant difference.

**C. What does this study add to what is already known?**

- This review revealed insufficient high-level evidence on the association of psychosocial factors at work and menstrual abnormalities.

## Introduction

Menstrual abnormalities are a crucial issue in women’s health. Menstrual cycle irregularities increased the risk of premature death,^1^ ovarian and breast cancer,^2,3^ cardiovascular disease,^4^ and diabetes.^5^ Moreover, fertility is essential for reproductive health and rights among preconceptual working-aged women. Menstrual characteristics and fertility are thus important indicators in occupational health settings.

Menstrual cycle and ovarian functions are regulated by sex hormones (i.e., the hypothalamic-pituitary-ovarian [HPO] axis); however, they are often disrupted by psychosocial factors^6,7^ through activation of the hypothalamic-pituitary-adrenal [HPA] axis.^8^ Psychosocial factors at work provide potential risks of activating the HPA axis and releasing cortisol due to psychological stress reactions, leading to menstrual disorders. For instance, some literature showed significant associations: low job control, co-worker support, and job security with high menstrual pain,^9^ low supervisor support with menopausal symptoms,^10^ strenuous work with premenstrual symptoms,^11,12^ work schedule variability with irregular cycles,^13,14^ time pressure and fast work speed with perimenstrual symptoms,^15^ and job strain (a combination of high job demands and low job control) with cycle length irregularities.^16^ The study design of many of them was cross-sectional, leading to limited evidence and suggestions for appropriate practices in the women’s occupational health field.

The HPA-HPO axis interaction is affected by circadian sleep control,^17,18^ The relation between night/shift work and menstrual disorders has been thus often described, and a meta-analysis by Stocker et al. (2014),^19^ which included a cross-sectional design, showed that shift work increased the risk of menstrual disruption and infertility^19^. However, more recent literature showed inconsistent results in the association of shift work with ovarian function.^20-22^ Lim et al. (2016) reported no association between shift work and ovarian functions or menstrual disorders in a cross-sectional design.^20^ In contrast, Minguez-Alarcon et al. (2017) reported that non-day work schedules were inversely related to oocyte production and quality among women receiving fertility treatment in a cross-sectional design.^21^ Only one longitudinal study by Albert-Sabater et al. (2016) reported that nurses on the rotating shift did not show an increased risk of having menstrual disorders compared with day staff.^22^ Since most studies were adopted a cross-sectional design (i.e., one cycle was treated as one individual outcome), the topics of shift work with menstruation require further investigations.

The association between psychosocial factors at work and menstrual abnormalities or fertility has not been systematically presented yet. Longitudinal studies should be summarized for concrete conclusions. The aim of this systematic review was thus to investigate whether psychosocial factors at work impact menstrual characteristics or fertility, limiting the literature that has utilized a longitudinal or prospective cohort design, assessing the outcome both at baseline and follow-up.

## Method

### Study design

The method was reported according to the Preferred Reporting Items for Systematic Review and Meta-Analysis (PRISMA 2020) guidelines.^23^ The study protocol has been registered at the UMIN registry (registration number: UMIN000039488 URL: https://upload.umin.ac.jp/cgi-bin/ctr/ctr_view_reg.cgi?recptno=R000044704). The registration date is 14th Feb 2020. The protocol paper has been available elsewhere.^24^

### PECO and Eligibility criteria

The eligible participants, exposures, comparisons, and outcomes (PECO) of this systematic review are listed in **Table 1**. PECO was: P: adult female workers (over 18 years old), E: adverse psychosocial factors at work (+), C: adverse psychosocial factors at work (−), and O: menstrual disorders, related symptoms, reproductive outcomes. The study design was restricted only longitudinal or prospective manner. The study was also excluded if the cross-sectional analysis method was applied, even if the study used a longitudinal design. The language was limited to English or Japanese.

**Table 1.** Eligibility criteria in the present systematic review.

|  | Inclusion criteria | Exclusion criteria |
| --- | --- | --- |
| Design | ● Longitudinal or prospective cohort design | ● Outcome was not assessed at least at two time point.<br>● Menstrual cycle was cross-sectionally analyzed as an independent unit. |
| Participants | ● Adult female workers (over 18 years old) | ● Pregnant workers<br>● Clinical students |
| Exposures | ● Presence of adverse psychosocial factors at work | N/A |
| Comparisons | ● Absence of adverse psychosocial factors at work | N/A |
| Outcomes | ● Menstrual disorders and related symptoms: 1) diverse menstrual dysfunctions (e.g., cycle disorder, hypermenorrhea), 2) gynecological disease/syndrome (e.g., endometriosis, polycystic ovarian syndrome), 3) menstrual-related symptoms (e.g., PMS, menopausal symptoms)<br>● Reproductive outcomes (e.g., infertility, time to be pregnant)<br>● Biological outcomes (e.g., serum sex hormone) | ● Pregnancy-related outcomes (e.g., premature birth)<br>● Malignant outcomes (e.g., cancer) |
| Article type | ● Original article published by peer-reviewed journal (including advanced online publication) | ● Conference abstract<br>● Letter/correspondence |
| Language | ● Written in English or Japanese | N/A |

### Search and Information Sources

Databases including PubMed (MEDLINE), Embase, PsycINFO, PsycARTICLES, and the Japan Medical Abstract Society database were searched for publications from inception to 26^th^ February 2020. The details of the search terms are shown in **Appendix 1**. The search terms for psychosocial factors at work were defined based on the previous meta-analysis.^25-28^ The search terms for outcome variables were initially selected by an investigator (NS) who reviewed previous systematic reviews handling menstrual-related health.^19,29-32^ The medical doctor in obstetrics and gynecology (outside the research members) confirmed the search terms before searching. The terms were amended to exclude infectious, inflammatory disease, and pelvic organ prolapse due to the lack of probability that psychosocial work-related factors cause these.

### Study selection

Microsoft Excel (Washington, USA) was used to manage all identified studies. An investigator (NS) excluded duplicate records. The remaining articles were shared with 16 investigators (NS, KI, KW, EM, HE, AI, KT, YH, YKom, MIi, YO, ASa, YA, MIw, YKob, and RI); Pairs of them independently assessed the title and abstract of each article according to the eligibility criteria (shifting phase: first screening). Subsequently, two investigators independently reviewed the full texts that were included in the first screening. Any disagreements were discussed until reaching a consensus by all authors. The reasons why studies were excluded were recorded during the full-text review phase.

### Data collection process and data items

Data were extracted independently from eligible articles by 16 investigators (NS, KI, KW, EM, HE, AI, KT, YH, YKom, MIi, YO, ASa, YA, MIw, YKob, and RI). An investigator (NS) integrated and formatted the information on publication year, study design, the country of study origin, the number of participants completing the baseline survey and included in the statistical analysis, demographic characteristics of the participants (i.e., age, occupation), the length of follow-up and attrition rate, exposure variables (i.e., adverse psychosocial factors at work), and outcome variables (i.e., menstrual abnormalities, fertility).

### Risk of Bias in individual Studies

The risk of bias in observational studies of exposures (ROBINS-I) tool was used to assess the quality of included studies. Investigators (NS, KI, KW, ASa, and YH) independently scored the bias classified as low, high, or unclear. Discrepancies in quality assessment among the investigators were solved by discussion and consensus among all authors. A Summary of Findings (SoF) will be created using the Grading of Recommendations, Assessment, Development and Evaluation (GRADE) approach to grade the certainty of evidence.

### Synthesis of Results and Meta-Analysis

For the main analysis, we synthesized all types of psychosocial factors at work and all types of menstrual-related disorders/symptoms. Meta-analysis was conducted if at least three eligible studies were found to have the same outcome. If a meta-analysis was not appropriate (i.e., only two or fewer studies were eligible and included), the results were presented in a narrative format.

### Changes to the protocol

To examine the association between exposure and outcome longitudinally, the study applied a cross-sectional fashioned analysis (i.e., one menstrual cycle was treated as an individual unit of outcome) was excluded.

## Result

### Database searching

Database searching yielded 12,868 abstracts (PubMed n = 2,823, EMBASE n = 9,340, PsycINFO n = 608, Japan Medical Abstracts Society n = 97). After removing 1,435 duplicates, 11,433 records were included in the first screening, after which 11,395 records were excluded, and 38 records proceeded to full-text screening. Subsequently, 32 studies that did not meet the criteria for design (n=13),^11-16,20-22,33-36^ participants (n=4),^37-40^ exposures (n=10),^41-50^ outcomes (n=2),^51,52^ article type (n=1),^53^ and duplicates (n=2) were excluded. Finally, six studies were included in the systematic qualitative review.^54-59^ The meta-analysis was not conducted due to insufficient studies based on our protocol. The flowchart of study selection is shown in **Figure 1**.

**Figure 1.**
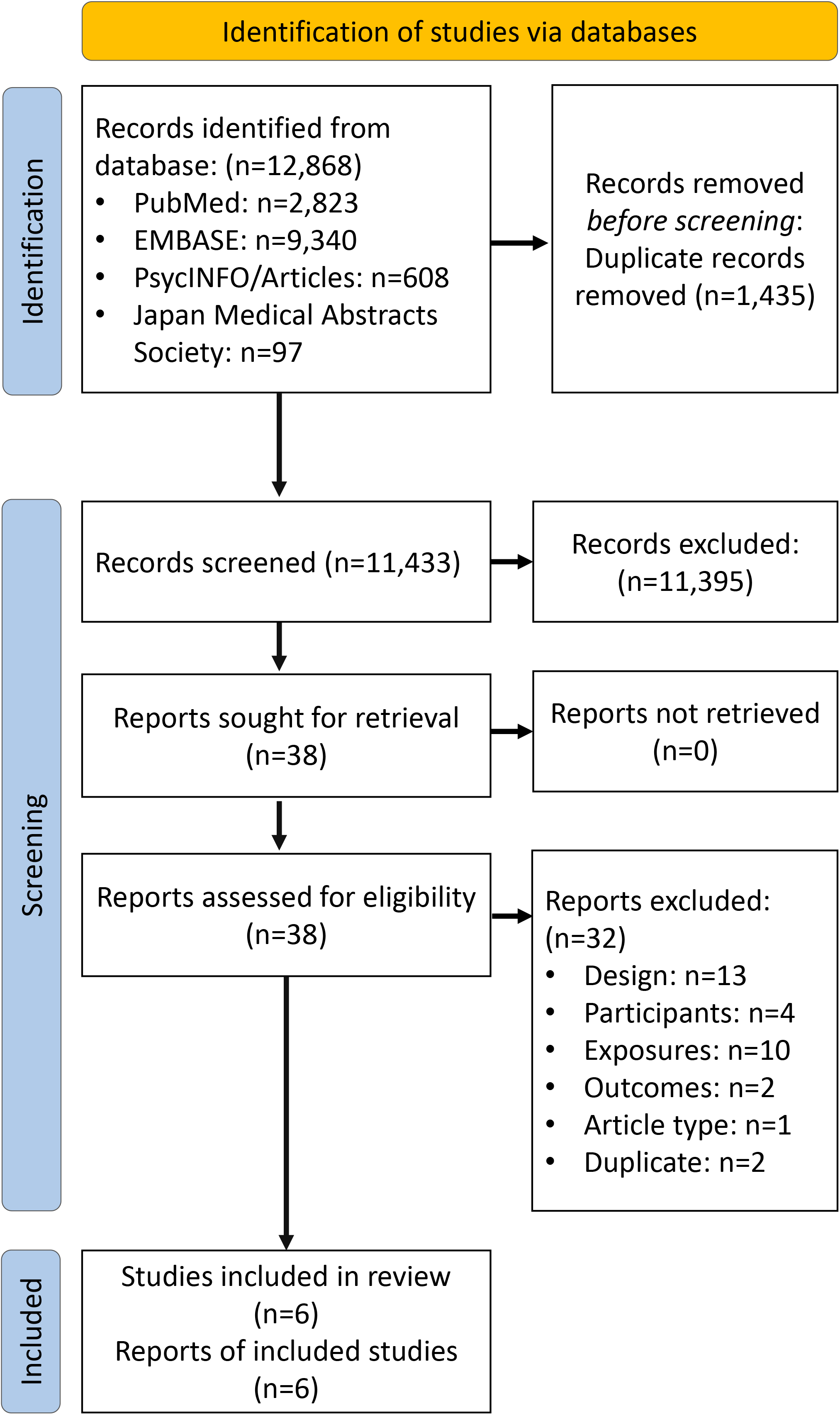
PRISMA 2020 Flow Diagram of systematic review search results.

### Study description

The characteristics of the six included studies are shown in **Table 2**. Three studies treated fertility outcomes.^55,57,59^ Earlier menopause,^56^ and incident diagnosis of endometriosis,^58^ and serum reproductive hormone^54^ were each measured by one study. No study was found which outcome was menstrual abnormalities. Three of the six studies retrieved their data from the nurse’s health study (NHS). All studies measured shift work as an adverse psychosocial factor at work. Each study reported job demands^59^ and long working hours. ^57^

**Table 2.**
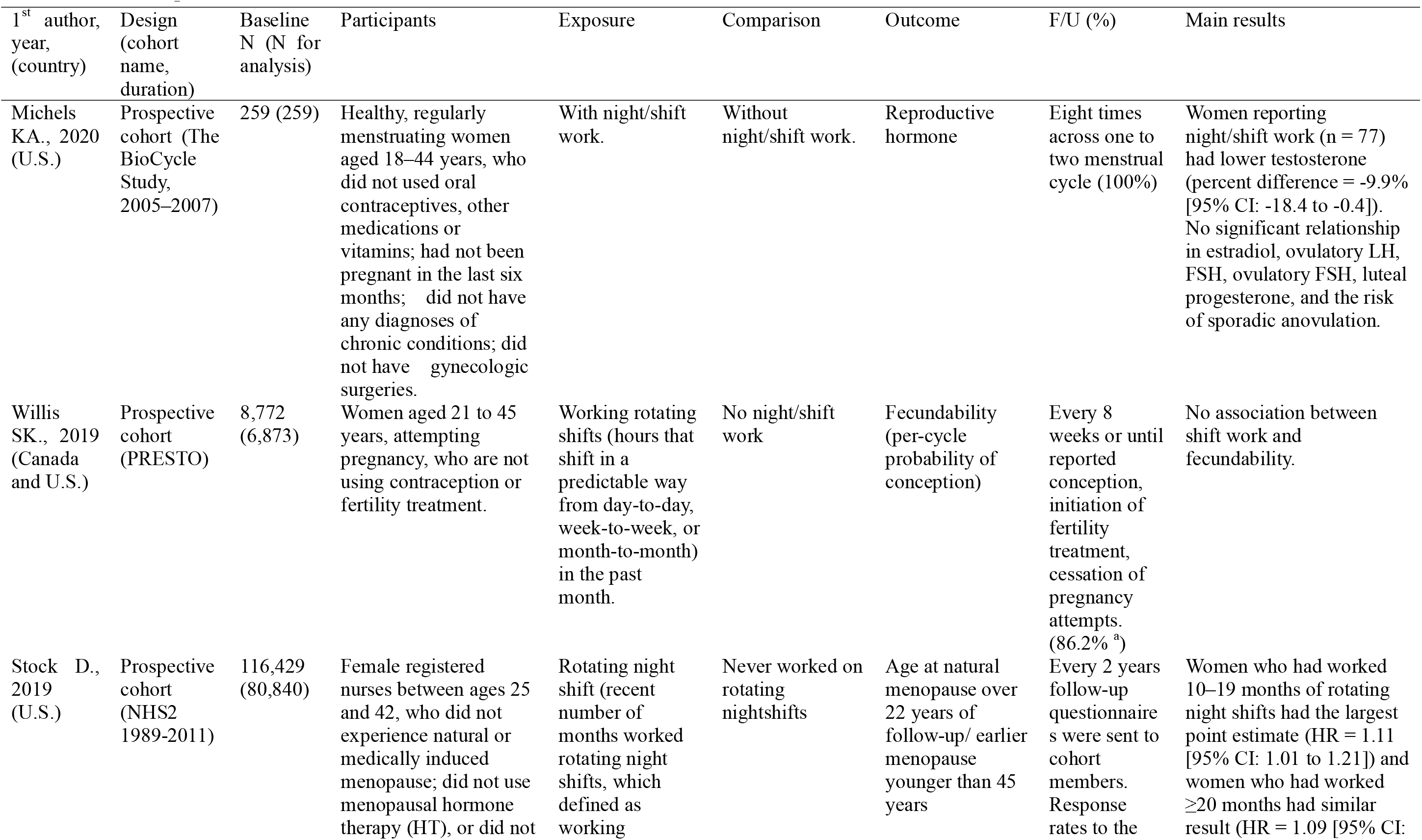
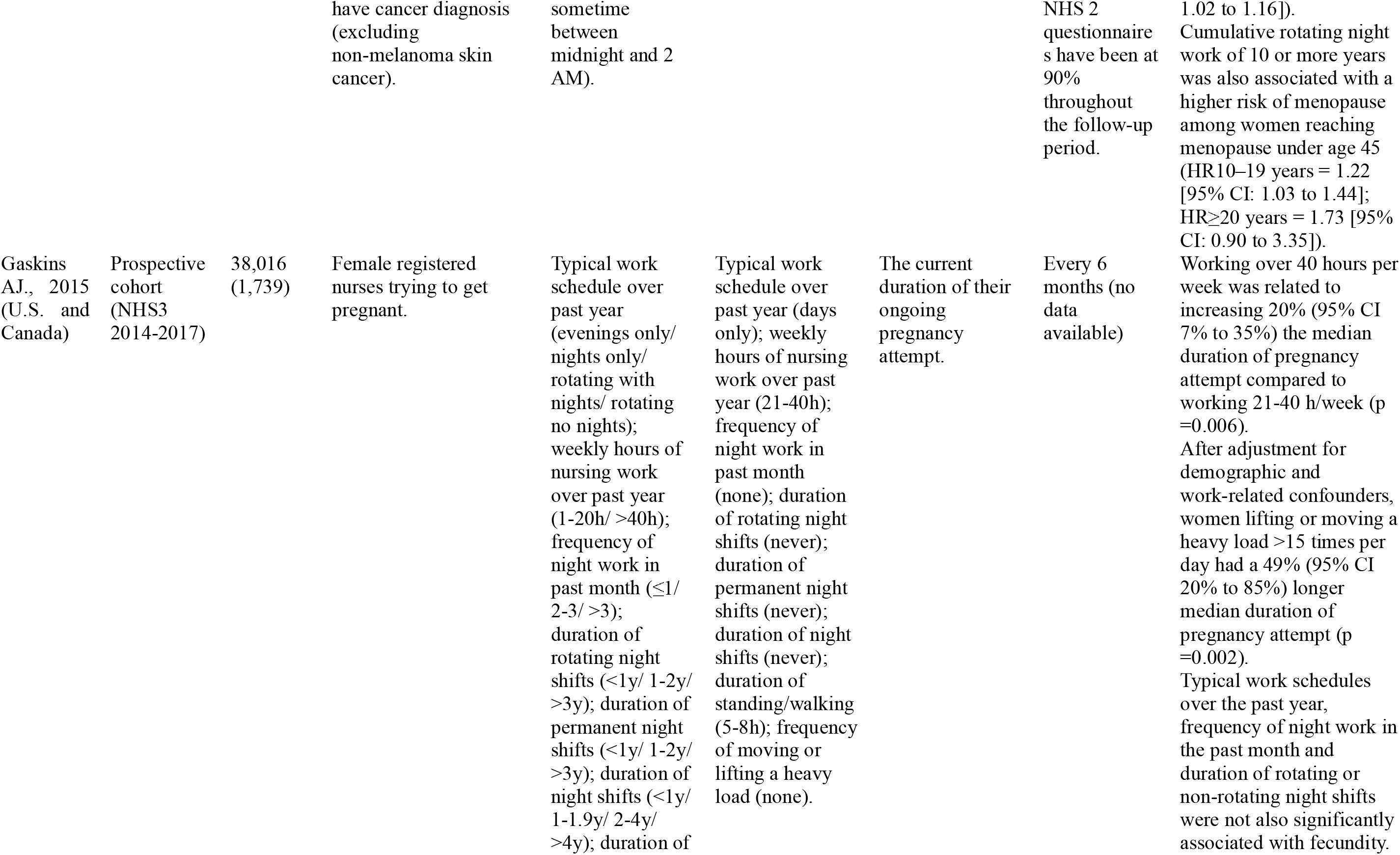
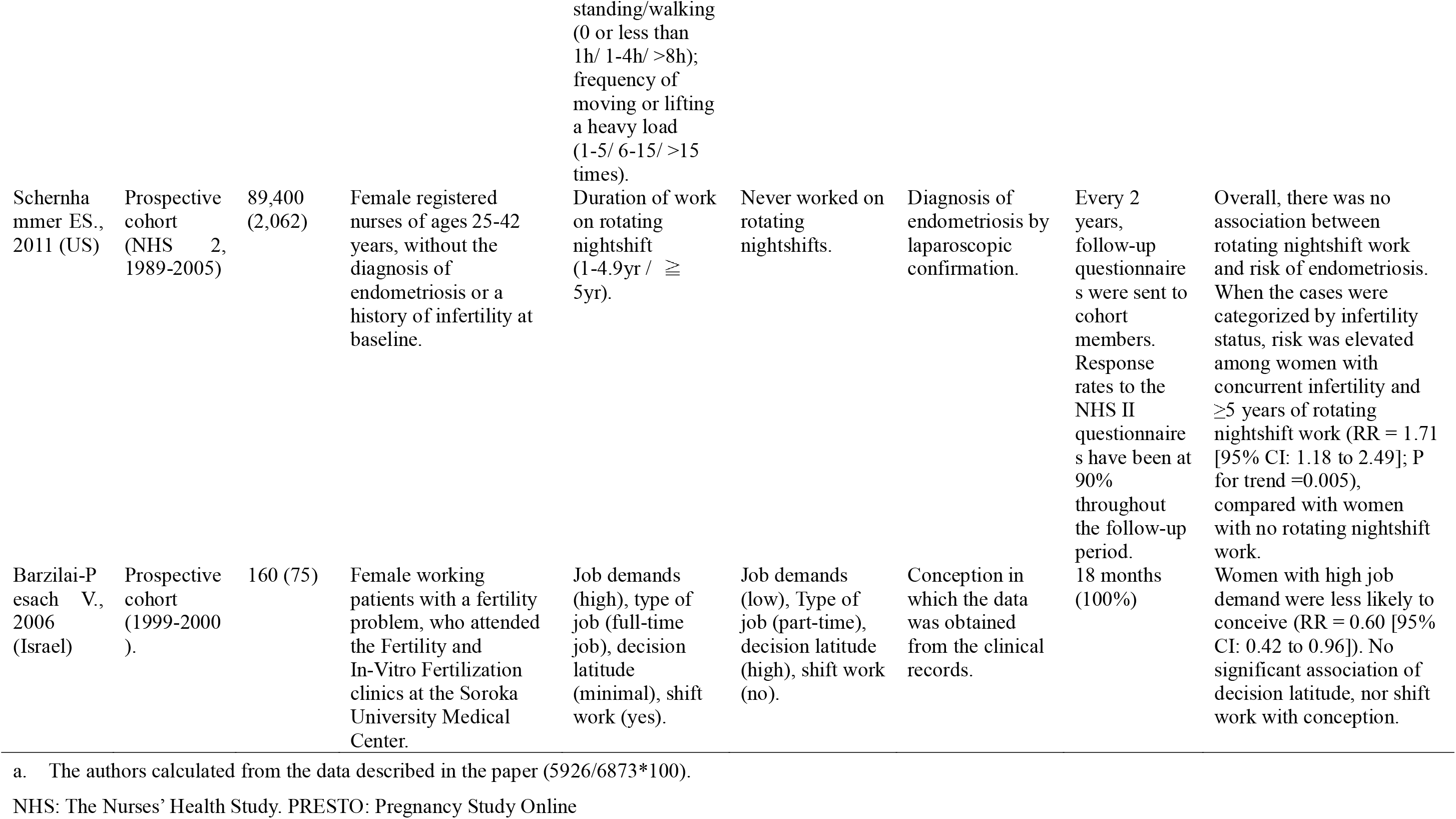
The descriptions of the included studies (N=6).

Although authors designed the protocol to conduct a meta-analysis if at least three eligible studies were found with the same outcome, fertility outcomes of included studies varied, and it was difficult to synthesize the findings: time to pregnancy,^55^ the current duration of participants’ ongoing pregnancy attempt,^57^ and conception.^59^

Regarding the three studies focusing on fertility outcomes, Barzilai-Pesach et al. (2006)^59^ prospectively evaluated the association between women’s occupational stress factors (i.e., job demands, decisional latitude, shift work, and working hours, measured using a self-reported questionnaire) and the outcome of fertility treatments. They followed 75 women (mean age = 31.3) who had worked at least three months before intake, had a female fertility problem and attended fertility clinics between 1999 and 2000. Information regarding the outcome of fertility treatments was based on a follow-up 18 months after the initial intake through fertility clinics. Women with high job demands were less likely to conceive (relative risk [RR] = 0.60 [95% confidence interval [CI]: 0.42 to 0.96]). There was no significant association of decision latitude, shift work, or working hours with conception. Gaskins et al. (2015) ^57^ evaluated the association of work schedule (i.e., work hours and total duration of night shift work) and physical factors (i.e., standing job and lifting load) with fecundity (i.e., current duration of participants’ ongoing pregnancy attempts) among female nurses, using the data from an internet-based cohort study of female nurses in the USA and Canada (NHS 3). For assessment of fecundity, the authors took a participant’s first report of ongoing pregnancy attempt after the baseline questionnaire as her outcome in three follow-up survey questionnaires conducted every six months. As a result, 1,739 women (median age = 33) were included in this analysis. Long working hours (over 40 hours per week) and lifting or moving a heavy load (>15 times per day) were related to a longer median duration of pregnancy attempts (increasing 20% [95% CI: 7% to 35%] and 49% [95% CI: 20% to 85%], respectively). Additionally, a subgroup analysis found that the association between heavy moving and lifting with reduced fecundity was even more pronounced among overweight or obese women. Night shift work over the past year was not significantly associated with fecundity. Willis et al. (2019)^55^ prospectively evaluated the association of shift work with fecundability. They followed North American women (living in Canada or the United States) who are not using contraception and aged 21–45 years attempting pregnancy for up to 12 months or until conception, using web-based surveys. Time to pregnancy was estimated by reports of their last menstrual period, usual menstrual cycle length, and the number of cycles they had attempted pregnancy at study entry. As a result, 6,873 women who had shift/night work ranging from 14.3% to 23.0% were included in the study. There was no association between shift work and fecundability.

Stock et al. (2019)^56^ prospectively evaluated the association between nurses’ rotating night shift work (i.e., the number of months with at least three nights per month in the past two years) and menopausal age, using the data from an ongoing prospective cohort study (NHS 2). This cohort included 116,429 female registered nurses in the United States, aged between 25 to 42, at the baseline survey in 1989, followed from 1991 through 2013. Participants were asked whether their menstrual periods had ceased, at what age, and if this was due to natural menopause, chemotherapy or radiation, or surgery. Of those whose menstrual periods had not stopped before the last follow-up, 27,456 had natural menopause. The mean age of natural menopause in these women was 50 (SD ±4.0) years old. Among them, 2,524 had menopause at under 45 years old. Significant associations were found between the number of months of night shift work in the past two years and earlier menopause (HRs = 1.06 to 1.11 among women with <10 months, 10-19 months, and with ≥20 months, compared to none of night shift work). Cumulative rotating night work of 10 or more years was also associated with a higher risk of menopause among women reaching menopause under age 45. These analyses did not consider any confounders such as psychological stress, job strain, or chronotype-related tolerance.

Schernhammer et al. (2011)^58^ reported an association between rotating night shift work and endometriosis risk within the NHS2. Participants were asked whether they had ever had physician-diagnosed endometriosis. Analyses of incident diagnosis of endometriosis were restricted to those women who reported laparoscopic confirmation of their diagnosis. This study included 2,062 laparoscopically confirmed cases documented during 16 years of follow-up evaluation from 89,400 women without diagnosed endometriosis at baseline. Overall, there was no association between rotating night shift work and endometriosis. When the cases were categorized by infertility status, the risk was elevated among women with concurrent infertility and ≥5 years of rotating night shift work (RR = 1.71 [95% CI: 1.18 to 2.49]; P for trend =0.005), compared with women with no rotating night shift work.

Michels et al. (2020)^54^ examined the influences of night/shift work on the serum reproductive hormones. The participants were recruited in the United States from the BioCycle Study (2005–2007), a prospective cohort of 259 healthy women (mean age = 24) not using oral contraceptives. At baseline, participants were asked if they were currently employed and if they worked nights or rotating shifts. Fasting blood samples were collected at eight clinic visits across each menstrual cycle. Fertility monitors were used to assist in the timing of mid-cycle clinic visits. Two cycles were followed. The proportion of participants who had night/shift work was 34.4%. The results showed that night/shift work did not influence estradiol or progesterone. Night/shift work showed lower testosterone (−9.9%) than employees without night/shift work, but the clinical implication of the finding was not mentioned.

### Risk of bias assessment

The summary of the risk of bias assessment by ROBINS-I is shown in **Table 3**. The overall judgment was all “serious” due to confounding or selection of reported results. The certainty of evidence was judged to be very low for all outcomes, downgraded for very serious risk of bias and publication bias (**Appendix 2**).

**Table 3.** Summary of risk of bias assessment for the included studies analyzed by ROBINS-I (N=6).

|  | Michels et al<br>(2020) #3722 | Willis et al.<br>(2019) #4093 | Stock et al.<br>(2019) #4307 | Gaskins et al.<br>(2015)#7240 | Schernhammer<br>et al.<br>(2011)#9737 | Barzilai-Pesach<br>et al.<br>(2006)#11436 |
| --- | --- | --- | --- | --- | --- | --- |
| 1. Confounding | S | S | M | S | S | S |
| 2. Selection of Participants | L | L | L | L | S | S |
| 3. Classification of intervention | M | M | M | M | S | S |
| 4. Deviations from intended interventions | L | L | L | L | L | L |
| 5. Missing data | S | M | S | L | S | S |
| 6. Measurement of outcomes | L | S | S | S | L | L |
| 7. Selection of reported result | S | M | S | S | S | S |
| Overall judgment | S | S | S | S | S | S |

Note: L: low risk of bias, M: moderate risk of bias, S: serious risk of bias, C: critical risk of bias, N: no information
ROBINS-I: The risk of bias in observational studies of exposures

## Discussion

### Principal Findings

This review investigated the association between psychosocial factors at work and menstrual abnormalities or fertility by limiting the study design to longitudinal, assessing the outcome at baseline and follow-up. Of the six included studies, three showed non-significant associations of shift work with fertility.^55,57,59^ Instead, high job demands, long working hours, and heavy lifting were significantly related.^57,59^ Rotating night shift work was not associated with the onset of endometriosis,^58^ but was with early menopause.^56^ One study identified slight changes in the sex hormones by shift work.^54^ All the studies had a serious risk of bias. There were still insufficient well-designed studies available to conduct meta-analysis and draw conclusions. However, the present study clarified the lack of evidence, achieved hypotheses to be examined in the future, and suggested further research directions.

### Comparison with Existing Literature

All three studies^55,57,59^ that investigated the risk of shift/night work for pregnancy outcomes showed non-significant associations. These studies were not included in the recent meta-analysis published in 2014^19^ that examined the influence of shift work on reproductive outcomes, showing that shift work increased the risk of infertility (crude odds ratio [OR]: 1.80 [95%CI: 1.01-3.20]). This meta-analysis included four cross-sectional fashioned studies (one study was a retrospective cohort) from 1996 to 2003.^60-63^ These studies retrieved a pregnant population and obtained the occupational data retrospectively. Three of them did not show significant associations,^60-62^ and one did.^63^ Certainly, some studies seemed to find that women with night/shift work tend to take a long time to become pregnant.^60-62^ However, the significance disappeared after adjusting the demographic and job characteristics information. The adjusted results of the meta-analysis also showed non-significant associations (adjusted OR: 1.12 [0.86 – 1.44]).^19^ From both our findings and previous reviews, we cannot conclude that shift/night work itself has an influence on fertility. More studies should be conducted using rigorous longitudinal study designs that account for possible confounding factors (psychosocial factors at work or lifestyle factors) to examine the association between shift work and fertility outcomes.

Two studies examined psychosocial factors at work other than shift/night work These studies showed that high job demands, long working hours, and heavy lifting were significantly associated with infertility.^57,59^ These findings were consistent with previous cross-sectional studies that were not included in the present review, which suggested that long working hours and physically demanding jobs lead to less ovarian function and infertility.^21,61^ In contrast, another study reported a non-significant association.^60^ Therefore, job demands, long time working, and lifting are possibly related to fertility outcomes. Although the previous study suggested that psychological stress reactions (i.e., depression, low positive affect) declined ovarian function,^64^ whether these psychosocial factors at work affect fertility through such distress or not (direct impact) still needs examination. Psychological stress theoretically disrupts HPA-HPO axis interactions ^8^ and has been associated with higher levels of oxidative stress, which potentially declines ovarian function. However, there is a lack of evidence to investigate the associations between psychosocial factors at work and fertility outcomes in a robust design. Further prospective studies will be expected in this area.

One study in this review showed the increased risk of early menopause from shift/night work.^56^ Rotating night shift work experience in the prior 2 years increased the risk of earlier menopause. The subgroup analysis with women under 45 showed that over 10 years of cumulative rotating night shift work exposure increased the risk of early menopause.^56^ This finding is consistent with the recent systematic review that included cross-sectional studies investigating job stress and menopause; the authors reported that age at menopause and severity of menopausal symptoms were both influenced by job-related factors such as high job strain. The review identified the risk factor for early menopause as smoking, type of work (e.g., service work), and shift work. Early menopause has been regarded as a significant outcome because it increases the risk of cardiovascular disease and all-cause mortality.^65^ Further investigations are needed to prevent the deterioration of female workers’ health if any occupational factors cumulatively affect early menopause. In this review, one study showed no significant association of rotating night shift work with the onset of endometriosis diagnosed by laparoscopy.^58^ Night shift work was found to decrease melatonin levels,^66^ reduced the aromatase activity and worked as a potential antiestrogenic agent. ^67^ Elevated estrogen levels caused by the low melatonin levels from night shift work could promote endometriosis growth. A population-based case-control study reported that any night shift work was associated with a 48% increased risk of endometriosis.^68^ The authors of the included study discussed the possible bias that women with clinical pain symptoms or with children might be likely to be out of the workforce in rotating work.

Additionally, laparoscopic confirmation of endometriosis among those who never experienced infertility is restricted to those with clinical symptoms, leading to underestimating the risk in those who never experienced infertility. The associations of shift work with endometriosis still need further investigation.

No study investigated the association of psychosocial factors at work with menstrual abnormality using longitudinal data. This review was limited to studies that evaluated the outcome (menstrual abnormality) in both baseline and follow-up. Furthermore, studies were excluded that analyzed the menstrual cycle as one unit of outcome. No study that met these criteria was found in this review. Some studies analyzed the cycle data as one unit.^14,16,22,34^ That method did not allow for a before/after comparison of data from one person and was considered a cross-sectional study. Menstrual abnormality can be caused not only by occupational stress but also by multiple other factors (e.g., body mass index [BMI], non-occupational stress). A longitudinal study evaluating the change in menstrual abnormalities will be needed to estimate the risk of psychosocial factors at work.

### Strengths and Limitations

This systematic review was the first review to investigate the evidence of the associations between psychosocial factors at work and menstrual abnormalities or fertility, which limited the longitudinal well-designed studies. However, this systematic review has several limitations. First, we did not conduct a meta-analysis due to an insufficient number of studies. The concrete evidence thus was not provided for the clinical questions. Second, this review is limited by English and Japanese language restriction; therefore, studies in other languages may have been missed. Third, publication bias may occur, but this review did not assess that bias.

### Conclusions and Implications

This systematic review investigated the association of psychosocial factors at work with menstrual abnormalities and fertility outcomes, and six articles were included. This review presented insufficient high-level evidence of well-designed studies on this topic. Because psychosocial factors at work are modifiable risk factors for women’s reproductive outcomes, further investments are needed to determine the implications for occupational health practice.

## Data Availability

All data produced in the present study are available upon reasonable request to the authors.

## Acknowledgments

The authors wish to thank Dr. Daisuke Shigemi (obstetrician-gynecologist) for a valuable suggestion on the variable selection.

We extend great thanks to cooperated researcher Yumi ASAI for her support of our procedures of screening the literature.

## Appendix 1 Search terms for PubMed

(employe*[tw] OR manag*[tw] OR colleague*[tw] OR worksit*[tw] OR “work”[tw] OR works*[tw] OR work’*[tw] OR worka*[tw] OR worke*[tw] OR workg*[tw] OR worki*[tw] OR workl*[tw] OR workp*[tw] OR occupant*[tw] OR company*[tw] OR offic*[tw] OR busines*[tw] OR workplace[mh]) AND ((“Stress, Mechanical”[Mesh] OR “Lifting”[Mesh] OR “Moving and Lifting Patients”[Mesh] OR “Weight-Bearing”[Mesh] OR “Biomechanics” OR “Physical Exertion”[Mesh] OR “Torsion, Mechanical”[Mesh] OR “Postural Balance”[Mesh] OR “Walking”[Mesh] OR “Recovery of Function”[Mesh] OR “Relaxation”[Mesh] OR (static[Title/Abstract] AND posture) OR (awkward[Title/Abstract] AND posture) OR (dynamic[Title/Abstract] AND posture) OR static work[Title/Abstract] OR dynamic load*[Title/Abstract] OR lift*[Title/Abstract] OR carry*[Title/Abstract] OR hold*[Title/Abstract] OR pull*[Title/Abstract] OR drag*[Title/Abstract] OR push*[Title/Abstract] OR manual handling[Title/Abstract] OR force*[Title/Abstract] OR biomechanic*[Title/Abstract] OR walking*[Title/Abstract] OR postural balance[Title/Abstract] OR flexion*[Title/Abstract] OR extension*[Title/Abstract] OR turning[Title/Abstract] OR sitting[Title/Abstract] OR kneeling[Title/Abstract] OR squatting[Title/Abstract] OR twisting[Title/Abstract] OR bending[Title/Abstract] OR reaching[Title/Abstract] OR standing[Title/Abstract] OR sedentary[Title/Abstract] OR repetitive movement*[Title/Abstract] OR monotonous work[Title/Abstract] OR relaxation[Title/Abstract] OR recovery of function[Title/Abstract] OR physical demand*[Title/Abstract] OR physically demand*[Title/Abstract]) OR (“Stress, Psychological”[Majr] OR “Social Support”[Majr] OR “Job Satisfaction”[Mesh] OR “Work Schedule Tolerance”[Mesh] OR “Employee Performance Appraisal”[Mesh] OR “Employee Grievances”[Mesh] OR “Social Justice/psychology”[Mesh] OR “Personnel Downsizing”[Mesh] OR “Staff Development”[Mesh] OR “Organizational Culture”[Mesh] OR “Bullying”[Mesh] OR “Prejudice”[Mesh] OR “Social Discrimination”[Mesh] OR “Interpersonal Relations”[Mesh] OR “Communication/psychology”[Mesh]) OR (psychosocial[Title/Abstract] OR job strain[Title/Abstract] OR work strain[Title/Abstract] OR work demand*[Title/Abstract] OR job demand*[Title/Abstract] OR high demand*[Title/Abstract] OR low control[Title/Abstract] OR lack of control[Title/Abstract] OR work control[Title/Abstract] OR job control[Title/Abstract] OR decision latitude[Title/Abstract] OR work influence*[Title/Abstract] OR demand resource*[Title/Abstract] OR effort reward*[Title/Abstract] OR time pressure*[Title/Abstract] OR recuperation*[Title/Abstract] OR work overload*[Title/Abstract] OR work over-load*[Title/Abstract] OR recovery[Title/Abstract] OR coping[Title/Abstract] OR work ability[Title/Abstract] OR social support[Title/Abstract] OR support system*[Title/Abstract] OR social network*[Title/Abstract] OR emotional support[Title/Abstract] OR interpersonal relation*[Title/Abstract] OR interaction*[Title/Abstract] OR social capital [Title/Abstract] OR justice*[Title/Abstract] OR injustice*[Title/Abstract] OR job satisfaction[Title/Abstract] OR work satisfaction[Title/Abstract] OR boredom[Title/Abstract] OR skill discretion*[Title/Abstract] OR staff development[Title/Abstract] OR discrimination[Title/Abstract] OR harass*[Title/Abstract] OR work-place conflict*[Title/Abstract] OR workplace violen*[Title/Abstract] OR work-place violen*[Title/Abstract] OR bullying[Title/Abstract] OR ageism[Title/Abstract] OR homophobia[Title/Abstract] OR racism[Title/Abstract] OR sexism[Title/Abstract] OR victimization*[Title/Abstract] OR silent workplace*[Title/Abstract] OR role ambiguity[Title/Abstract] OR role-conflict*[Title/Abstract] OR work-role*[Title/Abstract] OR working hour*[Title/Abstract] OR working time[Title/Abstract] OR day-time[Title/Abstract] OR night-time[Title/Abstract] OR shift work*[Title/Abstract] OR work shift*[Title/Abstract] OR temporary work[Title/Abstract] OR full-time[Title/Abstract] OR part-time[Title/Abstract] OR flexible work*[Title/Abstract] OR organizational change[Title/Abstract] OR organisational change[Title/Abstract] OR lean production[Title/Abstract] OR job security[Title/Abstract] OR job insecurity[Title/Abstract])) AND (menstruation OR menstrual OR “menstrual pain” OR “period pain” OR “period pains” OR menorrhagia OR hypermenorrhea OR dysmenorrhea[MeSH] OR premenstrual OR “PMS” OR “premenstrual syndrome” OR “PMDD” OR “premenstrual dysphoric disorder” OR amenorrhea OR menoxenia OR metrorrhagia OR endometrio* OR myoma* OR leiomyoma OR fibroids OR adenomyosis OR ovarian cyst* OR endometrioma OR polycystic ovar* OR menopaus* OR hot flush* OR climacteric OR estrogen* OR estradiol OR progesterone* OR prolactin OR lutenizing OR LH OR “lutenizing hormone” OR follicul* OR FSH OR “follicle stimulating hormone” OR gonadotropin OR GnRH OR “gonadotropin releasing hormone” OR “basal body temperature” OR infertility OR fertility OR fecund* OR subfecundity OR reproductiv*) AND (longitudinal OR prospective OR cohort OR (follow AND up) OR observational OR (case AND (control or cohort)) OR case-control OR case-cohort)

## Appendix 2: GRADE overview

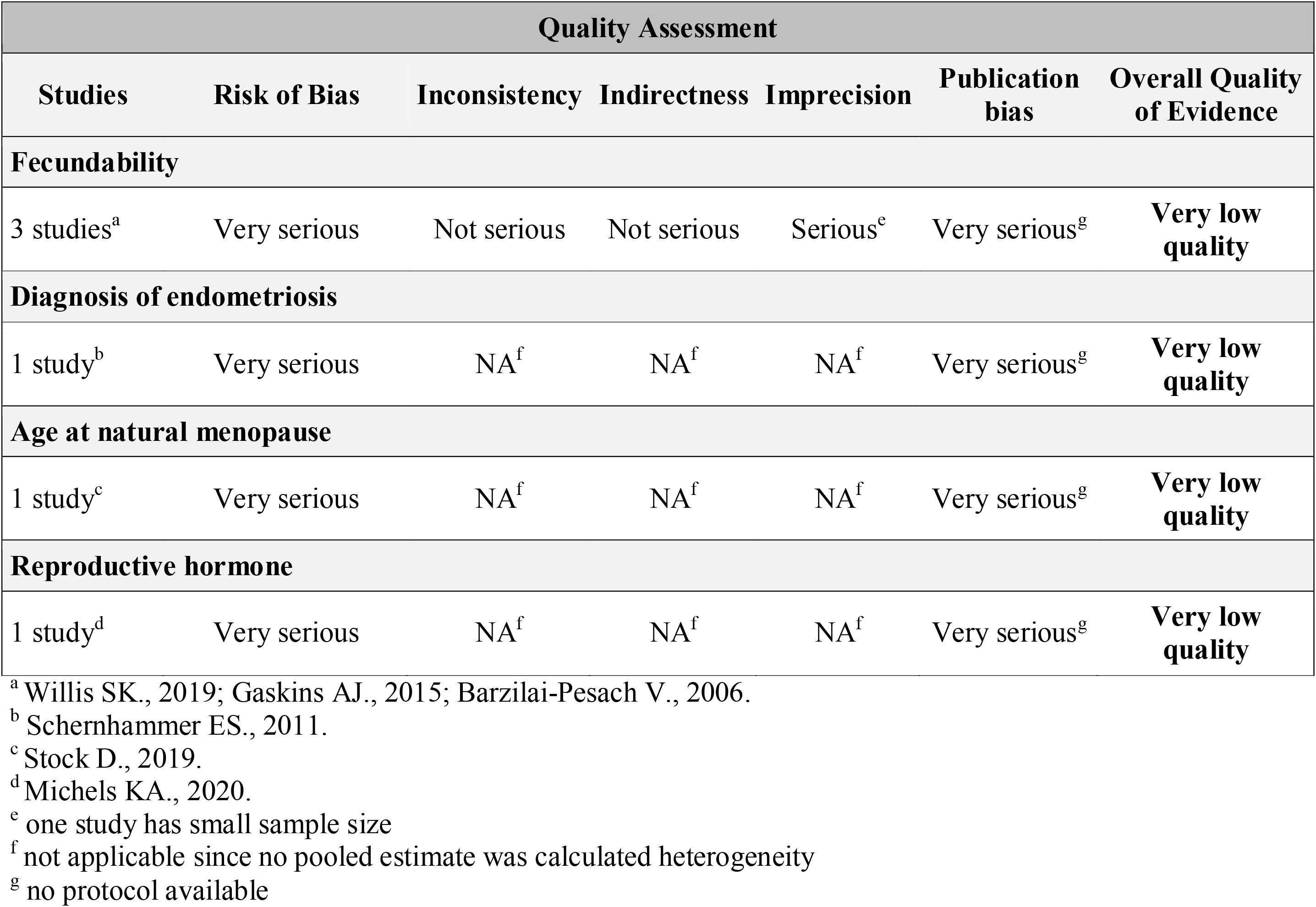

